# Analytical Validation of a Circulating Tumor DNA Assay using PhasED-Seq Technology for Detecting Residual Disease in B-Cell Malignancies

**DOI:** 10.1101/2024.08.09.24311742

**Authors:** Nina Boehm, Sandra Close, David M. Kurtz, Richard D. Hockett, Laura Hyland

## Abstract

**Background:** Circulating tumor DNA (ctDNA) is a non-invasive biomarker that can be used as a tool to detect minimal residual disease (MRD). MRD can provide important prognostic information in diffuse large B-cell lymphomas (DLBCL). Here, we present an MRD assay with an improved detection method for ctDNA, Phased Variant Enrichment and Detection Sequencing (PhasED-Seq) which leverages phased variants (PVs) to detect ctDNA.

**Methods:** Plasma samples from non-cancer controls were used to assess assay specificity. A limiting dilution series using DLBCL clinical-contrived samples was performed to assess assay sensitivity and precision. The accuracy of the PhasED-Seq-based assay was assessed using plasma samples from individuals with DLBCL and for whom MRD comparator assay results were also available. All samples were sourced from commercial vendors or academic studies.

**Results:** The analytical and clinical performance of the MRD assay was evaluated using clinical and clinical-contrived DLBCL samples. The assay’s false positive rate was 0.24% and the background error rate was 1.95E-08. The limit of detection at 95% detection rate (LoD95) at 120 ng was 0.7 parts in 1,000,000 and precision was >96%. Clinical accuracy was 90.62% PPA and 77.78% NPA.

**Conclusions:** The PhasED-Seq-based MRD assay has strong analytical and clinical performance in B-cell dyscrasias. Through the development of improved ctDNA detection methods such as that presented here, patient outcomes may be improved through the detection of residual disease or early relapse which may be used to guide treatment decisions.

**Brief Summary:** Here we present the analytical validation of a non-invasive minimal residual disease (MRD) assay which uses Phased Variant Enrichment and Detection Sequencing (PhasED-Seq) to improve the error profile and sensitivity of circulating tumor DNA (ctDNA) detection. The assay’s performance included a false positive rate of 0.24% and a background error rate of 1.95E-08. The limit of detection at 120 ng was 0.7 parts in 1,000,000 (6.61E-07 PVAF) with precision >96%. Positive and negative agreement were 90.62% and 77.78%, respectively. This suggests that the PhasED-Seq-based MRD assay is accurate and reproducible, thus appropriate for clinical use for individuals with B-cell malignancies.

## Introduction

Circulating tumor DNA (ctDNA), tumor DNA shed into the bloodstream, is a non-invasive biomarker that can be used as a tool to detect minimal residual disease (MRD). Detection of cancer-specific somatic mutations from ctDNA can provide clinically relevant information to predict therapeutic response, disease recurrence, and survival, and thus guide intervention decisions (1-3). As the utility of ctDNA detection has become appreciated, multiple investigational and commercially-available methods of detection have been developed (4). However, the sensitivity of first-generation approaches is limited and improved methods are needed to detect residual ctDNA when the tumor burden is low and the individual has a higher probability of responding to therapeutic intervention.

Diffuse large B-cell lymphoma (DLBCL) is the most common type of non-Hodgkin lymphoma (NHL) in the United States (5). Despite attempts to increase the efficacy of conventional first-line immunochemotherapy over the past two decades, approximately 40% of DLBCL patients still fail to respond or relapse (6). Current DLBCL response criteria rely on functional radiographic indices such as positron emission tomography/computed tomography (PET/CT) scans, which have limited sensitivity and specificity (7). There is a clear need to develop precision tools capable of rapid and accurate identification of patients harboring residual cancer burden who may be at high risk of relapse, such as the detection of residual tumor in the blood (i.e. ctDNA-MRD). We have developed a ctDNA-MRD platform based on Phased Variant Enrichment and Detection Sequencing (PhasED-Seq) to leverage phased variants (PVs) to improve the sensitivity of ctDNA detection (8). PVs are multiple somatic mutations in close proximity that can be concurrently observed on individual DNA molecules. PVs occur in most cancer types but are prevalent in stereotyped regions in B-cell malignancies (8), and are an attractive target to improve molecular detection techniques given their intrinsically low error profile (9). Here we describe the analytical validation of a sensitive PhasED-Seq-based MRD assay.

## Materials and Methods

### MRD Assay Overview

The Foresight CLARITY MRD assay (Foresight Diagnostics, Inc.) was assessed. This assay utilizes a tumor DNA sample (pre-treatment plasma or tumor tissue), a non-cancerous or normal DNA sample [e.g., peripheral blood mononuclear cell (PMBC) gDNA], and an MRD monitoring sample (plasma).

Extracted DNA from all samples is sequenced using a fixed hybrid capture panel (∼150kb) that enriches for genomic regions in areas that recurrently harbor PVs in B-cell lymphomas. Following sequencing, PVs are identified in the tumor and non-cancerous DNA samples to generate a tumor-specific somatic PV list. Tumor-specific PVs are defined as those that are present in the tumor DNA sample and absent in the normal DNA sample. This tumor-specific PV list is then used to assess for MRD in the MRD monitoring sample using informative molecules, or any cell-free DNA (cfDNA) molecules spanning the location of a tumor-specific PV that could harbor a PV (Figure 1). Mutant molecules are informative molecules containing the mutant allele. MRD is defined as the presence of tumor specific PVs (mutant molecules), meeting a threshold based on the likelihood of an incidental mutation overlapping with the tumor-specific PV list.

**Figure 1.**
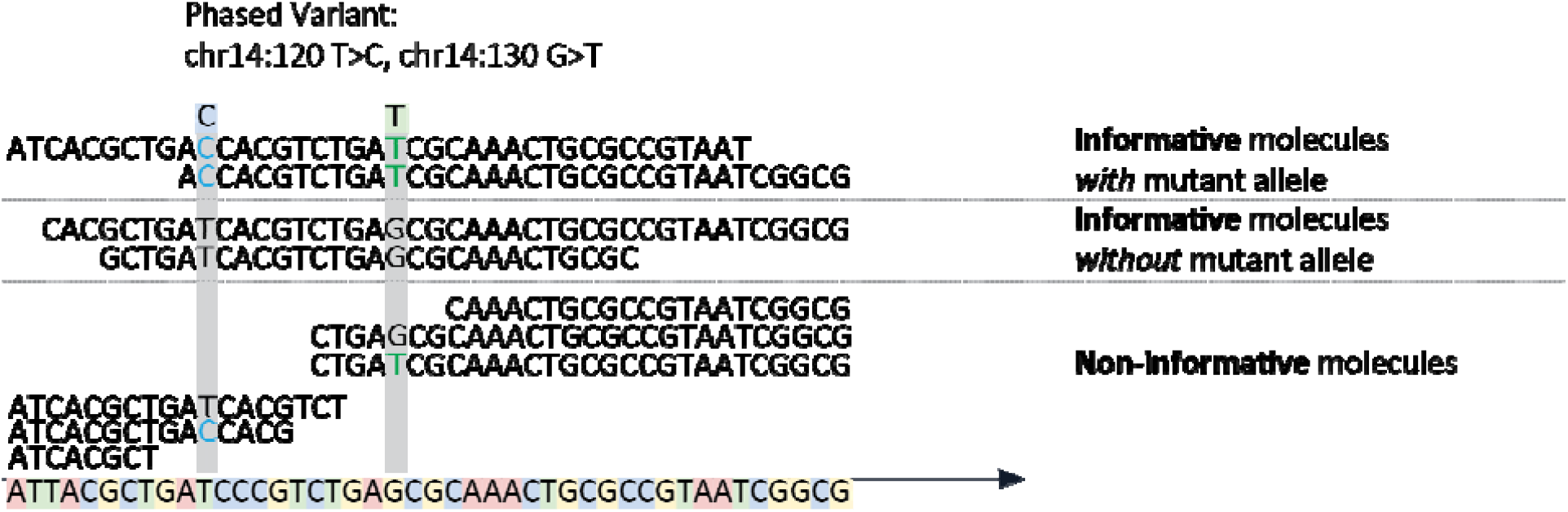
Depiction of Informative Molecules. ‘Informative Molecules’ are cfDNA molecules spanning the location of a tumor-specific PV. Any cfDNA molecules that could harbor a PV from the patient’s PV list are considered Informative Molecules. ‘Mutant Molecules’ are Informative Molecules which harbor the tumor-specific mutant allele of one or more PVs in the patient’s PV List. In this example the PV chr14:120T>C, chr14:130G>T spans 4 informative molecules and 2 mutant molecules.

### Sample Preparation

The analytical performance of the PhasED-Seq-based MRD assay was assessed in the Clinical Laboratory Improvement Amendments (CLIA)-registered laboratory at Foresight Diagnostics, Inc., following standard operating procedures. Samples included healthy donor samples (self-reported cancer-free at time of collection; N=169), clinical DLBCL samples (N=76), and clinical-contrived DLBCL samples (N=2). Clinical DLBCL samples were samples obtained from individuals with an active diagnosis of DLBCL. Clinical-contrived DLBCL samples were prepared by combining extracted cell-free DNA (cfDNA) from clinical samples and healthy donor samples; multiple clinical DLBCL and healthy donor samples were pooled to make clinical-contrived samples. The clinical-contrived samples were then diluted to a targeted phased variant allele fraction (PVAF). PVAF is defined as the ratio of molecules containing a tumor-specific PV per molecules spanning the positions of ≥1 PV. Both clinical and clinical-contrived samples were required to meet the following criteria for study inclusion: minimum input mass of 5 ng and ≥85% Phred quality score of 30 (Q30) from the Illumina sequencer. Sequencing metrics are reported within each study.

Both commercially procured (Discovery Life Sciences and BioIVT) and residual samples from academic research collaborations were utilized in the analytical validation studies. Samples from academic collaborations were collected with appropriate patient consent which allowed for research use of residual samples and institutional review board (IRB) oversight. Positive and negative controls were used along with each study sample batch. The positive control was a mix of lymphoma cell lines rich in PVs and the negative control consisted of libraries prepared with 50 µL nuclease-free water and carried through the entire workflow.

### DNA Isolation

cfDNA was isolated from plasma using the QIAsymphony DSP Circulating DNA Kit (Qiagen, Hilden, Germany; Catalog Number: 937556) on the automated QIAsymphony system. Double-stranded (dsDNA) was quantified by fluorometry using a Qubit Fluorometer with the Qubit dsDNA High Sensitivity Assay Kit (Invitrogen, Waltham, MA; Catalog Number: Q32854). gDNA was isolated from plasma-depleted whole blood (PDWB) or PBMCs using the commercial QIAsymphony DSP DNA Mini Kit (Qiagen, Hilden, Germany; Catalog Number: 937236) on the automated QIAsymphony system and sheared using sonication. DNA was quantified using the Qubit dsDNA Broad Range Assay Kit (Invitrogen, Waltham, MA; Catalog Number: Q32853).

### Library Preparation and Next-Generation Sequencing

Library preparation, hybrid capture target enrichment, and sequencing by synthesis was performed according to Foresight Diagnostics, Inc., optimized workflows under standard operating procedures. Five to 120 ng of cfDNA or gDNA were used to construct sequencing libraries using KAPA HyperPrep Kits (Roche Sequencing Solutions, Indianapolis, IN) on manual and automated custom workflows. Library DNA was enriched using a custom B-cell lymphoma probe panel (Integrated DNA Technologies, Inc.), performed per the manufacturer’s instructions using both a manual and an automated workflow on the Bravo Automated Liquid Handling platform. Following enrichment, libraries were sequenced using sequencing by synthesis on the Illumina NovaSeq 6000 and/or NovaSeq X Plus instrument (Illumina, San Diego, CA).

### Analysis of Sequencing Data and MRD Status Determination

Sequence data were analyzed using in-house developed algorithms and pipelines. Briefly, raw sequencing data were demultiplexed to FASTQ files for each sample using BCL Convert software (Illumina, San Diego, CA; Versions 2.2.0 to 2.4.0). Low-quality sequencing reads and adapter read-through were removed using fastp (version 0.20.0). Sequencing reads were then aligned to the reference genome (GRCh37) using BWA-MEM aligner (version 2.2.1) to create one alignment file per sample, followed by proprietary methods to remove polymerase chain reaction (PCR) and optical duplicates. The resulting sequence alignment file was used for the analysis of PVs. MRD status was determined by the presence or absence of tumor-specific PVs, meeting a threshold based on the likelihood of an incidental mutation overlapping with the tumor-specific PV list.

### Analytical Specificity

The assay specificity or limit of blank (LoB) was evaluated according to CLSI guidance EP17-A2 (10). EP17-A2 defines the LoB as the highest value expected to be observed from a series of measurements on a sample that contains no analyte (blank samples). Whole blood from 60 cancer-free donors (blank samples) was collected in Streck cfDNA blood collection tubes (BCTs; Streck, Catalog Number: 230470), processed to plasma, and cfDNA. DNA input mass into library preparation was 120 ng. Two library replicates were prepared from each donor and 120 libraries were generated for sequencing. Libraries were interrogated by DLBCL tumor-specific PV lists resulting in a MRD positive or negative call. The false positive rate (FPR) and background error rate for the assay was calculated per donor and overall.

### Analytical Sensitivity

To determine the limit of detection (LoD) of the MRD assay (95% detection rate per CLSI EP17-A2), a limited dilution series of a DLBCL clinical-contrived sample was prepared at 6 targeted PVAF levels (7.00E-06, 3.50E-06, 1.75E-06, 8.75E-07, 4.38E-07, and 2.19E-07). Clinical-contrived sample replicates were created by combining cfDNA from 4 DLBCL patient samples and diluting the mixture into background cfDNA from healthy donor plasma. Ten replicates were tested across 2 reagent lots.

Corrected targeted PVAF levels, based on the observed PVAFs, were used for detection rate and probit models. The detection rate for each level was calculated by adding the number of MRD positive calls and dividing the number by the total number of replicates tested. The probit model was used to compute the number of mutant molecules and PVAF corresponding to a detection rate of 95% for the sample. Probit model fit was acceptable by evaluating with a statistical goodness of fit test.

### Reproducibility and Repeatability

Assay precision was evaluated with a clinical-contrived MRD positive sample that was prepared across different targeted PVAF levels. At 120 ng input mass, targeted PVAF levels were 7.00E-06, 3.50E-06, and 1.75E-06 and at 5 ng input mass targeted PVAF levels were 0.0001, 0.00004, and 0.00002.

Average positive agreement (APA) was used to calculate the assay’s repeatability and reproducibility as described in Yu et al, 2016. (11). Sample replicates were prepared across 2 operators, 2 reagent lots, and 3 time points.

### Accuracy

The accuracy of the PhasED-Seq-based MRD assay was determined by comparing the results to a previously established single nucleotide variant (SNV)-based orthogonal method for detection of ctDNA using samples from individuals with DLBCL (12). Samples from a total of 19 individuals with DLBCL were utilized, including 19 pre-treatment plasma samples, 19 normal samples, 31 timepoint-of-interest plasma samples from timepoints of interest during treatment (Cycle 2, Day 1 or Cycle 3, Day 1) or at end-of-therapy (EOT). Results from the SNV-based MRD assay and PhasED-Seq-based MRD assay were compared. Discordant results between the two assays were adjudicated by comparison with clinical outcomes.

## Results

### Quality Control (QC) Pass Rate

The assay QC pass rate across all pre-analytic, analytic, and post-analytic metrics during the conduct of this analytical validation was 99.0%.

### Analytical Specificity

The assay specificity was assessed using cfDNA from 60 cancer-free donors (blank samples). All sample replicates passed QC metrics for 120 libraries for evaluation. Samples were sequenced to an average median depth of 21,321x and on-target coverage was >91%. As blank samples do not have tumor-derived PVs, PV lists from 35 DLBCL patients were used to measure the FPR and background error rate. The 35 PV lists covered 65.5% of the total B-cell capture panel spanning multiple chromosomes. Each blank sample replicate was interrogated by 35 patient PV lists resulting in 4,200 possible tumor detection calls to evaluate the assay FPR (Table 1). The overall assay FPR was 0.24%. The background error rate of the PhasED-Seq-based MRD assay was 1.95E-08, or 1.95 mutant molecules in 100 million informative molecules.

**Table 1.**
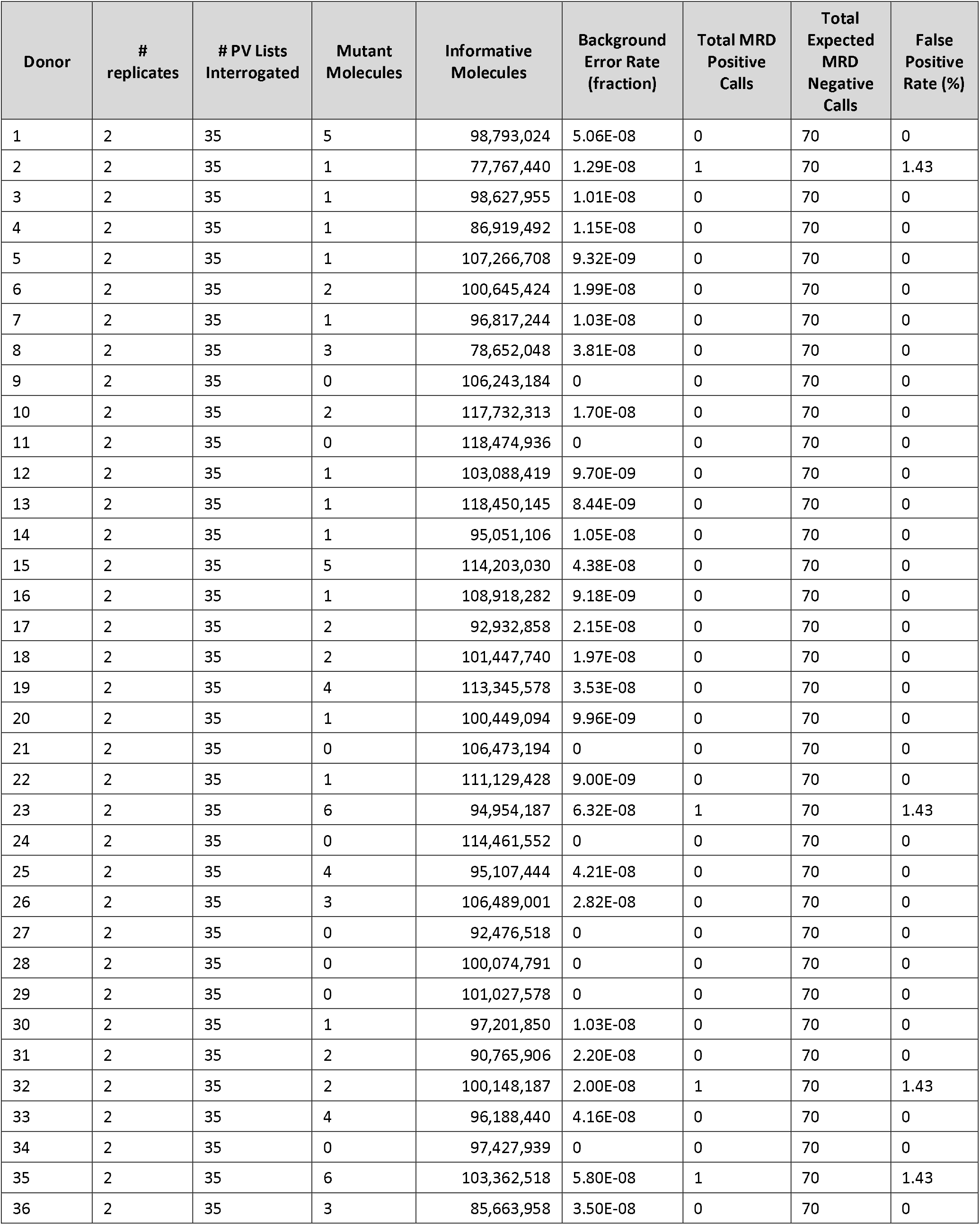
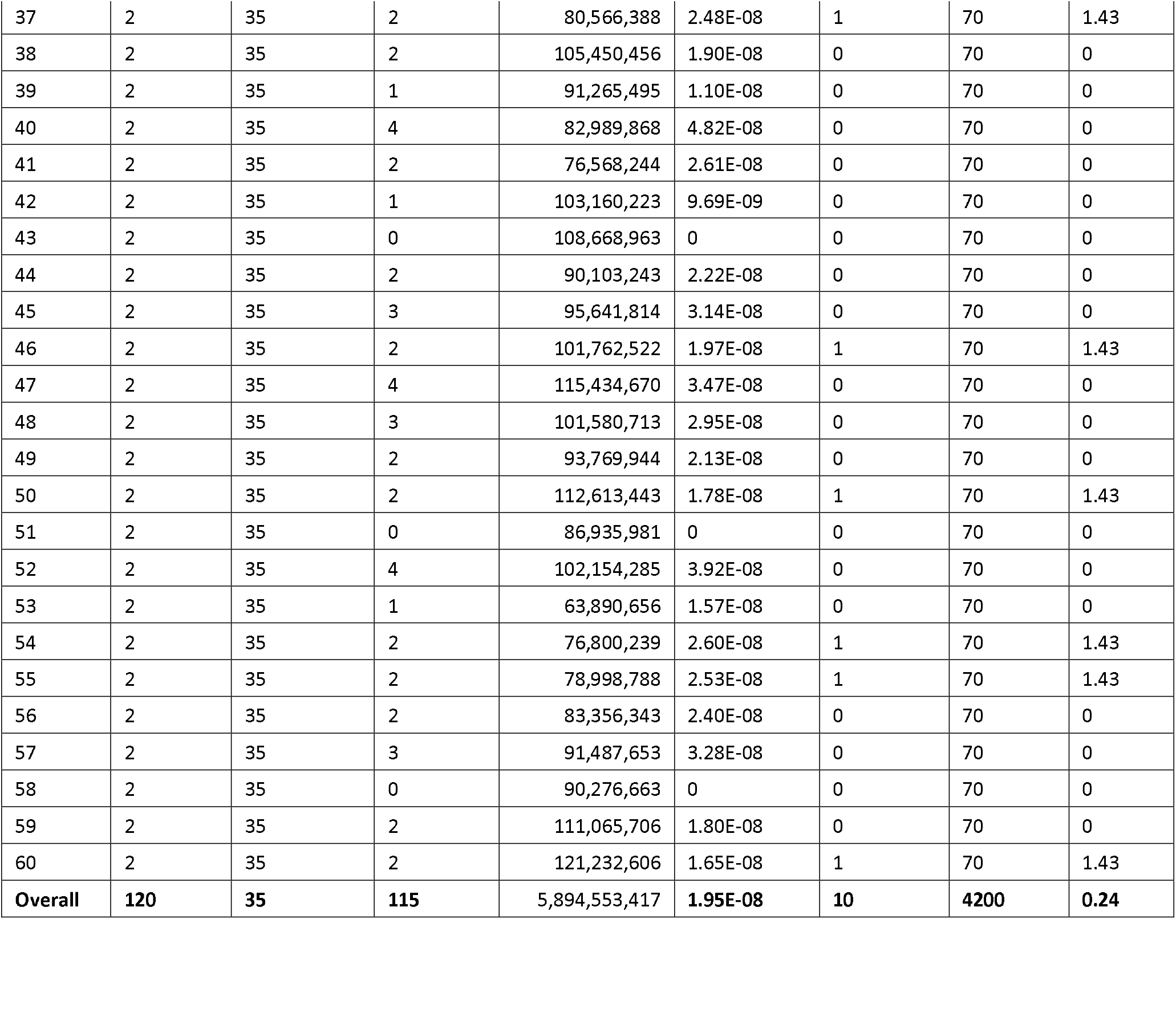
Overall Background Error Rate.

| Donor | # replicates | # PV Lists Interrogated | Mutant Molecules | Informative Molecules | Background Error Rate (fraction) | Total MRD Positive Calls | Total Expected MRD Negative Calls | False Positive Rate (%) |
| --- | --- | --- | --- | --- | --- | --- | --- | --- |
| 1 | 2 | 35 | 5 | 98,793,024 | 5.06E-08 | 0 | 70 | 0 |
| 2 | 2 | 35 | 1 | 77,767,440 | 1.29E-08 | 1 | 70 | 1.43 |
| 3 | 2 | 35 | 1 | 98,627,955 | 1.01E-08 | 0 | 70 | 0 |
| 4 | 2 | 35 | 1 | 86,919,492 | 1.15E-08 | 0 | 70 | 0 |
| 5 | 2 | 35 | 1 | 107,266,708 | 9.32E-09 | 0 | 70 | 0 |
| 6 | 2 | 35 | 2 | 100,645,424 | 1.99E-08 | 0 | 70 | 0 |
| 7 | 2 | 35 | 1 | 96,817,244 | 1.03E-08 | 0 | 70 | 0 |
| 8 | 2 | 35 | 3 | 78,652,048 | 3.81E-08 | 0 | 70 | 0 |
| 9 | 2 | 35 | 0 | 106,243,184 | 0 | 0 | 70 | 0 |
| 10 | 2 | 35 | 2 | 117,732,313 | 1.70E-08 | 0 | 70 | 0 |
| 11 | 2 | 35 | 0 | 118,474,936 | 0 | 0 | 70 | 0 |
| 12 | 2 | 35 | 1 | 103,088,419 | 9.70E-09 | 0 | 70 | 0 |
| 13 | 2 | 35 | 1 | 118,450,145 | 8.44E-09 | 0 | 70 | 0 |
| 14 | 2 | 35 | 1 | 95,051,106 | 1.05E-08 | 0 | 70 | 0 |
| 15 | 2 | 35 | 5 | 114,203,030 | 4.38E-08 | 0 | 70 | 0 |
| 16 | 2 | 35 | 1 | 108,918,282 | 9.18E-09 | 0 | 70 | 0 |
| 17 | 2 | 35 | 2 | 92,932,858 | 2.15E-08 | 0 | 70 | 0 |
| 18 | 2 | 35 | 2 | 101,447,740 | 1.97E-08 | 0 | 70 | 0 |
| 19 | 2 | 35 | 4 | 113,345,578 | 3.53E-08 | 0 | 70 | 0 |
| 20 | 2 | 35 | 1 | 100,449,094 | 9.96E-09 | 0 | 70 | 0 |
| 21 | 2 | 35 | 0 | 106,473,194 | 0 | 0 | 70 | 0 |
| 22 | 2 | 35 | 1 | 111,129,428 | 9.00E-09 | 0 | 70 | 0 |
| 23 | 2 | 35 | 6 | 94,954,187 | 6.32E-08 | 1 | 70 | 1.43 |
| 24 | 2 | 35 | 0 | 114,461,552 | 0 | 0 | 70 | 0 |
| 25 | 2 | 35 | 4 | 95,107,444 | 4.21E-08 | 0 | 70 | 0 |
| 26 | 2 | 35 | 3 | 106,489,001 | 2.82E-08 | 0 | 70 | 0 |
| 27 | 2 | 35 | 0 | 92,476,518 | 0 | 0 | 70 | 0 |
| 28 | 2 | 35 | 0 | 100,074,791 | 0 | 0 | 70 | 0 |
| 29 | 2 | 35 | 0 | 101,027,578 | 0 | 0 | 70 | 0 |
| 30 | 2 | 35 | 1 | 97,201,850 | 1.03E-08 | 0 | 70 | 0 |
| 31 | 2 | 35 | 2 | 90,765,906 | 2.20E-08 | 0 | 70 | 0 |
| 32 | 2 | 35 | 2 | 100,148,187 | 2.00E-08 | 1 | 70 | 1.43 |
| 33 | 2 | 35 | 4 | 96,188,440 | 4.16E-08 | 0 | 70 | 0 |
| 34 | 2 | 35 | 0 | 97,427,939 | 0 | 0 | 70 | 0 |
| 35 | 2 | 35 | 6 | 103,362,518 | 5.80E-08 | 1 | 70 | 1.43 |
| 36 | 2 | 35 | 3 | 85,663,958 | 3.50E-08 | 0 | 70 | 0 |
| 37 | 2 | 35 | 2 | 80,566,388 | 2.48E-08 | 1 | 70 | 1.43 |
| 38 | 2 | 35 | 2 | 105,450,456 | 1.90E-08 | 0 | 70 | 0 |
| 39 | 2 | 35 | 1 | 91,265,495 | 1.10E-08 | 0 | 70 | 0 |
| 40 | 2 | 35 | 4 | 82,989,868 | 4.82E-08 | 0 | 70 | 0 |
| 41 | 2 | 35 | 2 | 76,568,244 | 2.61E-08 | 0 | 70 | 0 |
| 42 | 2 | 35 | 1 | 103,160,223 | 9.69E-09 | 0 | 70 | 0 |
| 43 | 2 | 35 | 0 | 108,668,963 | 0 | 0 | 70 | 0 |
| 44 | 2 | 35 | 2 | 90,103,243 | 2.22E-08 | 0 | 70 | 0 |
| 45 | 2 | 35 | 3 | 95,641,814 | 3.14E-08 | 0 | 70 | 0 |
| 46 | 2 | 35 | 2 | 101,762,522 | 1.97E-08 | 1 | 70 | 1.43 |
| 47 | 2 | 35 | 4 | 115,434,670 | 3.47E-08 | 0 | 70 | 0 |
| 48 | 2 | 35 | 3 | 101,580,713 | 2.95E-08 | 0 | 70 | 0 |
| 49 | 2 | 35 | 2 | 93,769,944 | 2.13E-08 | 0 | 70 | 0 |
| 50 | 2 | 35 | 2 | 112,613,443 | 1.78E-08 | 1 | 70 | 1.43 |
| 51 | 2 | 35 | 0 | 86,935,981 | 0 | 0 | 70 | 0 |
| 52 | 2 | 35 | 4 | 102,154,285 | 3.92E-08 | 0 | 70 | 0 |
| 53 | 2 | 35 | 1 | 63,890,656 | 1.57E-08 | 0 | 70 | 0 |
| 54 | 2 | 35 | 2 | 76,800,239 | 2.60E-08 | 1 | 70 | 1.43 |
| 55 | 2 | 35 | 2 | 78,998,788 | 2.53E-08 | 1 | 70 | 1.43 |
| 56 | 2 | 35 | 2 | 83,356,343 | 2.40E-08 | 0 | 70 | 0 |
| 57 | 2 | 35 | 3 | 91,487,653 | 3.28E-08 | 0 | 70 | 0 |
| 58 | 2 | 35 | 0 | 90,276,663 | 0 | 0 | 70 | 0 |
| 59 | 2 | 35 | 2 | 111,065,706 | 1.80E-08 | 0 | 70 | 0 |
| 60 | 2 | 35 | 2 | 121,232,606 | 1.65E-08 | 1 | 70 | 1.43 |
| **Overall** | **120** | **35** | **115** | 5,894,553,417 | **1.95E-08** | **10** | **4200** | **0.24** |

### Analytical Sensitivity

DLBCL clinical-contrived sample replicates were prepared at 6 targeted PVAF levels. All replicates passed quality control metrics. Replicates were sequenced to an average median depth of 19,455x. The PV list generated for the clinical-contrived sample had a total of 9,043 PVs. In a dilution series ranging from 4.83E-06 to 1.51E-07, PVAF was linear with the dilution of mutant molecules (Supplementary Figure 1). Detection rates at each PVAF level are presented in Table 2. Based on probit modeling the mutant molecules and PVAF corresponding to a detection rate of 95%, the LoD of the MRD assay, is 3.11 mutant molecules and 6.61E-07 (Supplementary Figure 2), or 0.7 parts in 1,000,000.

**Table 2.** Analytical Sensitivity.

|  | Corrected PVAF Level | n | N | Detection Rate (%) |
| --- | --- | --- | --- | --- |
|  | 4.83E-06 | 10 | 10 | 100 |
|  | 2.42E-06 | 10 | 10 | 100 |
|  | 1.21E-06 | 10 | 10 | 100 |
|  | 6.04E-07 | 9 | 10 | 90 |
|  | 3.02E-07 | 5 | 10 | 50 |
|  | 1.51E-07 | 2 | 10 | 20 |
|  | 0 | 0 | 4 | 0 |
| LoD95 PVAF - Probit | 6.61E-07 |  |  |  |

PVAF, phased variant allele fraction; N, total replicates at level; n, MRD positive results; Multiple lots were used to evaluate the detection rates at each target PVAF level.

### Reproducibility and Repeatability

Table 3 shows the assay precision using clinical-contrived samples, covering 5 ng and 120 ng DNA input mass and the low to high analytical measurement range. All sample replicates across operators, reagent lots, and time points (N=104) passed QC metrics. Replicates were sequenced to an average median depth of 21,965x. Assay repeatability and reproducibility was >96% (Table 3).

**Table 3.**
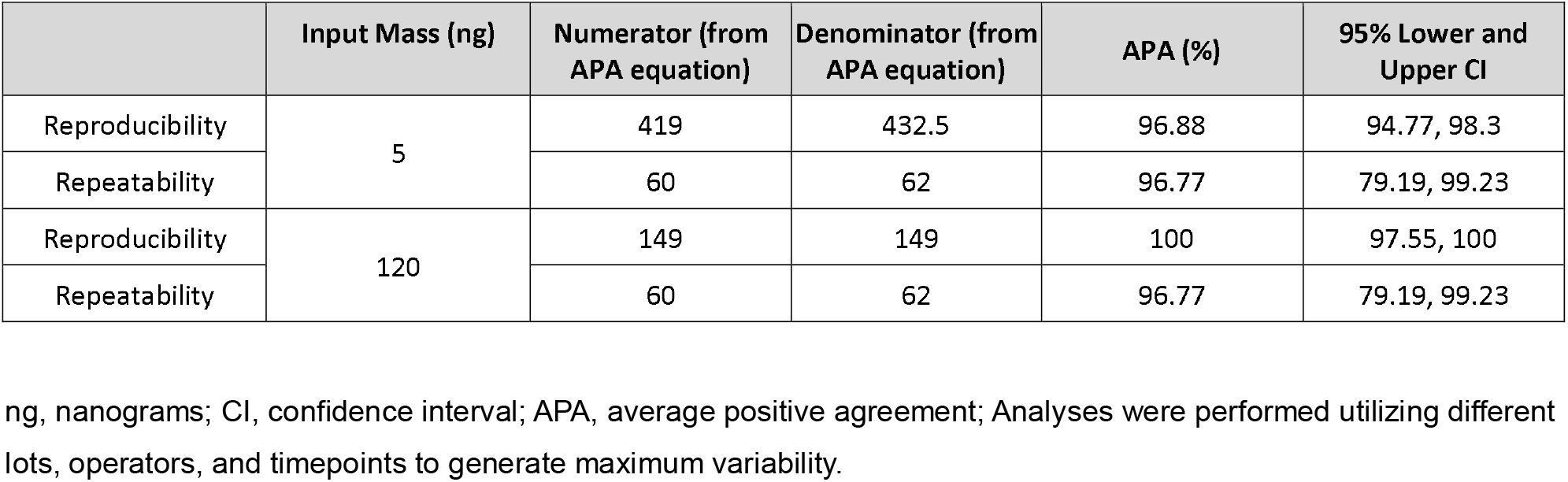
DLBCL MRD Assay Precision Results.

|  | Input Mass (ng) | Numerator (from<br>APA equation) | Denominator (from<br>APA equation) | APA (%) | 95% Lower and<br>Upper CI |
| --- | --- | --- | --- | --- | --- |
| Reproducibility | 5 | 419 | 432.5 | 96.88 | 94.77, 98.3 |
| Repeatability |  | 60 | 62 | 96.77 | 79.19, 99.23 |
| Reproducibility | 120 | 149 | 149 | 100 | 97.55, 100 |
| Repeatability |  | 60 | 62 | 96.77 | 79.19, 99.23 |

ng, nanograms; CI, confidence interval; APA, average positive agreement; Analyses were performed utilizing different lots, operators, and timepoints to generate maximum variability.

### Accuracy

Fifty samples were evaluated for concordance between the PhasED-Seq-based MRD assay and a previously established SNV-based method for MRD detection (12). All samples passed QC metrics. Library input mass for plasma cfDNA samples for MRD detection ranged from 21.3 to 80 ng and while non-cancerous, normal samples were prepared at 80 ng. Normal samples were sequenced to an average median depth of 4012x and cfDNA samples 5980x. The number of phased variants for each sample and donor was calculated and ranged from 1 to 1,816 PVs.

According to the comparator SNV-based method, 18 samples were called MRD negative and 32 were called MRD positive (Table 4). MRD monitoring samples that were called MRD positive by the comparator assay had a range of tumor fractions from 0.000022 to 0.1697. Using the PhasED-Seq-based MRD assay, 17 samples were called MRD negative and 33 samples MRD positive. The MRD positive samples as determined by the PhasED-Seq-based MRD assay had a range of PVAFs from 0.0000088 to 0.2567.

**Table 4.**
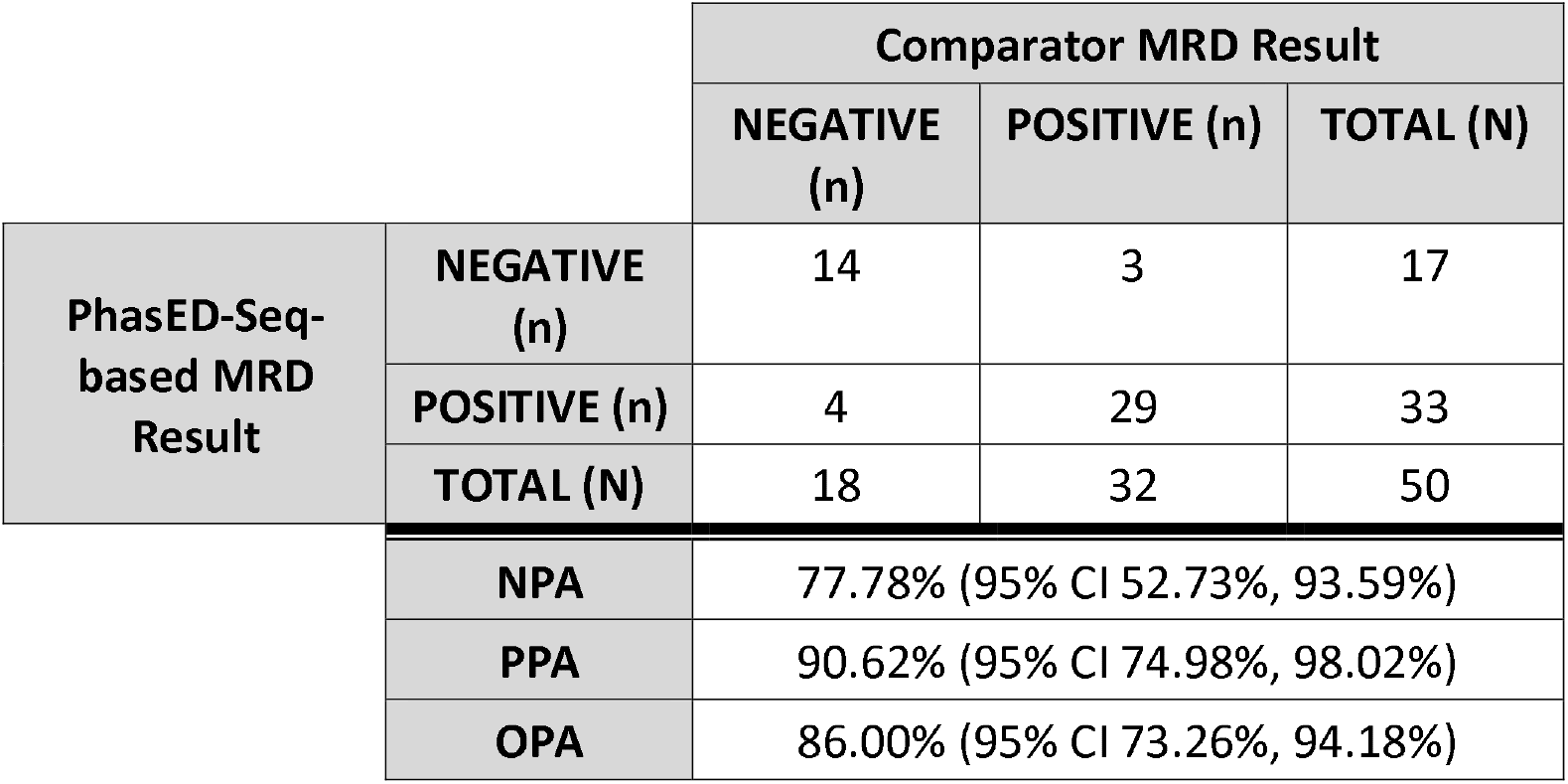
Accuracy Between-Test Concordance.

Positive percent agreement (PPA) for the MRD assay was 90.62% (95% CI 74.98%, 98.02%) and NPA was 77.78% (95% CI 52.73, 93.59; Table 4) using the SNV-based method as reference. There were 7 discordant calls between the two methods. In all the cases where the two assays were discordant (Table 5) the PhasED-Seq-based MRD assay agreed with the clinical outcomes (PPA 100%, NPA 100%). In the same cases, the SNV-based comparator assay had a lower concordance with clinical outcomes (PPA 0%, NPA 60%).

**Table 5.** Discordant Calls.

| Donor | Monitoring Timepoint | PhasED-Seq-based MRD Result | Comparator MRD Result | Clinical Outcome | MRD Call Discordance Explanation |
| --- | --- | --- | --- | --- | --- |
| P1001 | Baseline | NEGATIVE | POSITIVE | Disease Free | Patient cured; MRD assay result matches clinical outcome. MRD assay result at EOT was MRD NEGATIVE. |
| P1001 | Cycle2Day1 | NEGATIVE | POSITIVE | Disease Free | Patient cured; MRD assay result matches clinical outcome. MRD assay result at EOT was MRD NEGATIVE. |
| P1002 | Cycle2Day1 | POSITIVE | NEGATIVE | Disease Free | Blood collection was during treatment. Disease was still detected at Cycle2Day1, based on MRD assay result. Patient had additional cycles of therapy that likely cleared their disease. MRD assay result at Cycle3Day1 was MRD NEGATIVE which matches clinical outcome. |
| P1003 | Cycle2Day1 | POSITIVE | NEGATIVE | Disease Free | Blood collection was during treatment. Disease was still detected at Cycle2Day1, based on MRD assay result. Patient had additional cycles of therapy that likely cleared their disease. MRD assay result at EOT was MRD NEGATIVE matching the clinical outcome. |
| P1004 | End Of Therapy | NEGATIVE | POSITIVE | Disease Free | Patient cured with >5 years follow up; MRD assay result at EOT matches clinical outcome. |
| P1005 | Cycle3Day1 | POSITIVE | NEGATIVE | Disease Free | Blood collection was during treatment. Disease was still detected at Cycle3Day1, based on MRD assay result. Patient had additional cycles of therapy that likely cleared their disease. MRD assay result at EOT was MRD NEGATIVE which matches clinical outcome. |
| P1006 | Cycle3Day1 | POSITIVE | NEGATIVE | Disease Relapse | Patient still has disease based on MRD assay result. MRD assay result at EOT was MRD POSITIVE which matches clinical |

## Discussion

Here we describe the analytical validation of an MRD assay that utilizes PhasED-Seq technology to identify tumor-specific PVs to detect ctDNA in B-cell dyscrasias. The main benefit of using PVs for MRD detection is improving the signal-to-noise ratio in sequencing data by requiring the concordant detection of at least 2 separate non-reference events in an individual DNA molecule. Leveraging multiple somatic mutations within individual cfDNA fragments to detect ctDNA reduces the background error rate, as previously described (9). The use of multiple variants on the same DNA strand (i.e. PVs) provides an advantage at low ctDNA levels in which the specificity of SNV-based technologies is reduced due to the inherent background error rate of SNVs.

The analyses presented here demonstrate the analytical performance of this PhasED-Seq-based MRD assay. The high specificity of the MRD assay was demonstrated through analysis of samples from individuals without cancer (N=60), with a FPR of 0.24% and a background error rate of 1.95E-08, which is ∼1000-fold lower than reported for SNP-based technologies, even when utilizing unique molecular identifiers (9). With this low background error rate, the analytical sensitivity of the PhasED-Seq-based MRD assay was determined to be 0.7 part per 1 million (6.61E-07 PVAF). It is important to note that this analytical sensitivity was determined in the context of a limited amount of DNA input. To achieve this sensitivity, an adequate plasma sample providing >1,000,000 informative molecules must be utilized; therefore, starting from a larger amount of plasma or cfDNA is recommended. If an available sample does not contain sufficient cfDNA, the number of informative molecules will dictate the level at which ctDNA can be detected. Indeed, even the LoD study described here is affected by the number of informative molecules and DNA input and, with higher amounts of DNA input, an improved LoD would be expected.

Taken together, the assay’s high sensitivity and specificity suggests a reliable assay to detect low PVAFs without the accumulation of false-positive signal. At the increased sensitivity level, the assay’s reproducibility and repeatability rate was >96% and proved to be robust to operator, reagent lot, and timepoint variability. Comparison of this PhasED-Seq-based MRD assay against an orthogonal SNV-based approach for detecting ctDNA-MRD demonstrated a high overall concordance. The discordant calls were adjudicated against patient clinical outcome data and for all discordant cases, the PhasED-Seq-based MRD assay correlated with clinical outcome.

Collectively, the data presented here suggests that the PhasED-Seq-based MRD assay is accurate and reproducible, making it appropriate for use in the clinical setting for individuals with B-cell malignancies. Through the development of improved ctDNA detection methods such as that presented here, patient outcomes may be improved through the detection of residual disease or early relapse which may be used to guide treatment decisions.

## Data Availability

All data produced in the present study are available upon reasonable request to the authors.

## Acknowledgements

The authors would like to acknowledge Krystal Brown, PhD and Stephanie Meek, PhD, for their assistance in preparation of this manuscript.

