## Supplementary Figure 1 for "Analytical Validation of a Circulating Tumor DNA Assay using PhasED-Seq Technology for Detecting Residual Disease in B-Cell Malignancies"

### Supplementary Figure 1. Limiting Dilution Series Linearity

Corrected phased variant allele frequency (PVAF) for each dilution level of the limit of detection (LoD) clinical-contrived sample were determined by using the average of the observed PVAF for the first dilution level and then using the dilution factors performed in the series. For example, if the second dilution was a 1:2 dilution, then the average PVAF from the first dilution was divided by 2. The observed PVAF at these targets were plotted. Red triangles represent average of observed PVAF at the corrected PVAF level. If observed and corrected PVAF aligned, the red triangles would produce a line with a slope of 1. The dotted line in each figure represents a slope of 1 (i.e, x=y) and is shown for visualization.

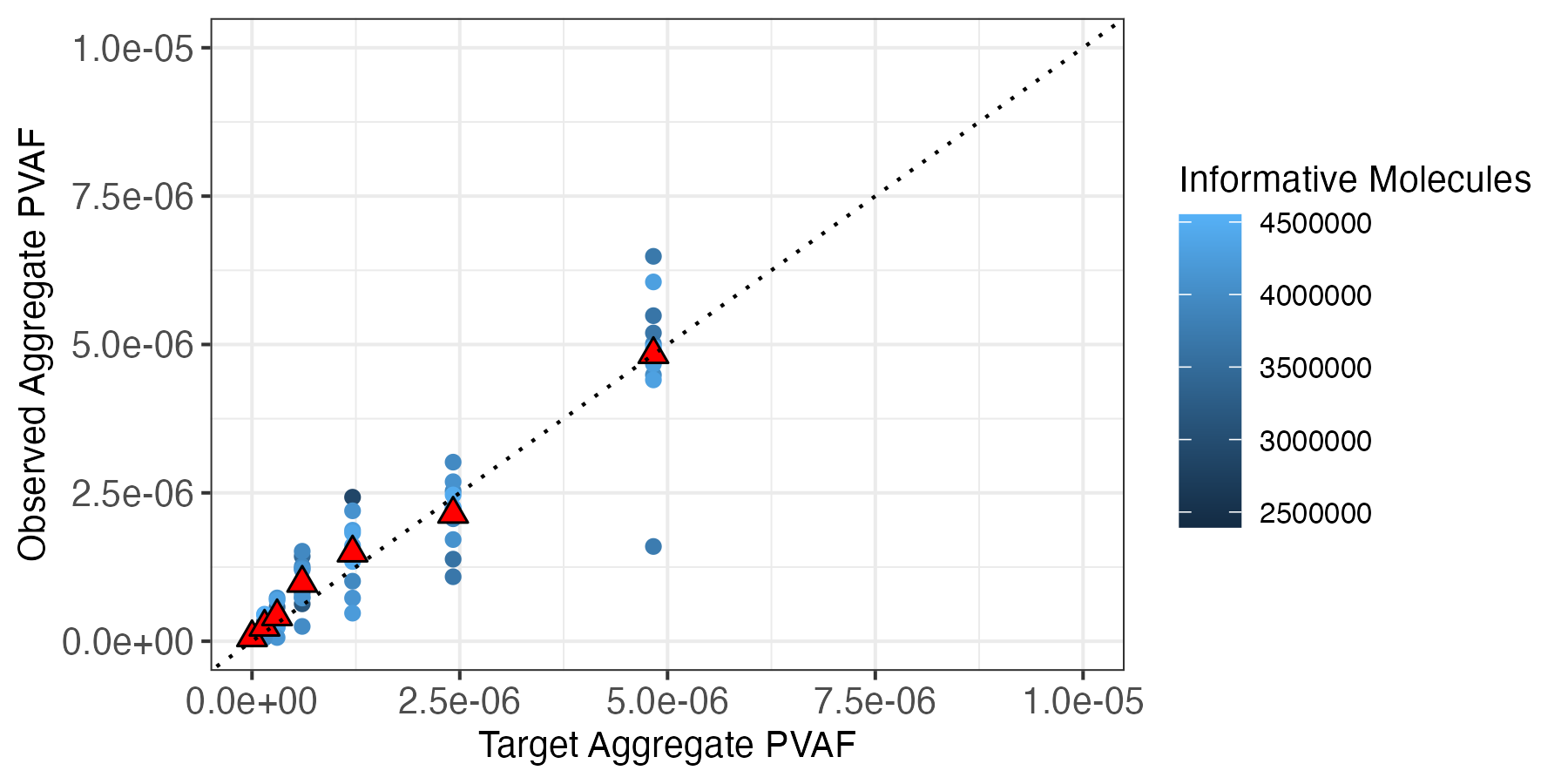
