## Supplementary Figure 2 for "Analytical Validation of a Circulating Tumor DNA Assay using PhasED-Seq Technology for Detecting Residual Disease in B-Cell Malignancies"

### Supplementary Figure 2. PROBIT Graph

The relationship between detection rate and PVAF is modeled by probit model regression. The black dots represent the observed data points while the solid black line represents probit model’s predict probabilities. The detection rates at the specific PVAF levels are presented as the number of MRD positive calls/total number of replicates. The dotted line represents the detection rate at 0.95 (95%). Blue text is the PVAF at 95% detection rate for the sample or the limit of detection (LoD).

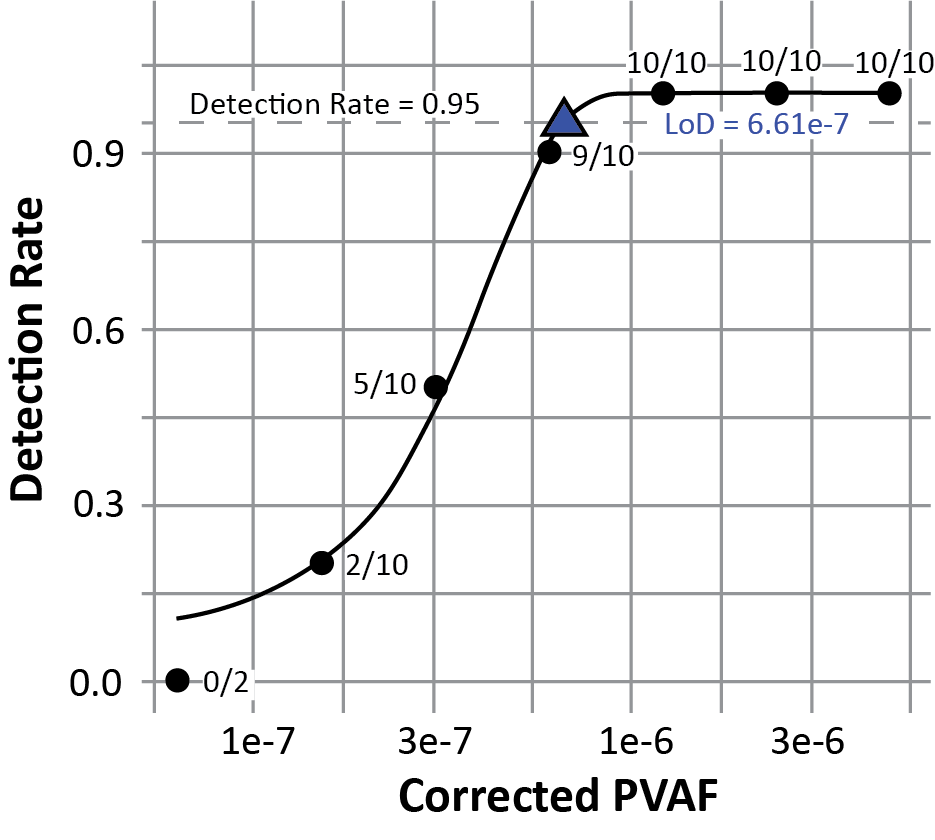
